# Extracellular microvesicle microRNAs, along with imaging metrics, improve detection of aggressive prostate cancer

**DOI:** 10.1101/2024.08.23.24312491

**Authors:** Kapil K Avasthi, Jung Choi, Tetiana Glushko, Brandon J Manley, Alice Yu, Julio Pow-Sang, Robert Gatenby, Liang Wang, Yoganand Balagurunathan

## Abstract

Prostate cancer is the most commonly diagnosed cancer in men worldwide. Early diagnosis of the disease provides better treatment options for these patients. Magnetic resonance imaging (MRI) provides an overall assessment of prostate disease. Quantitative metrics (radiomics) from the MRI provide a better evaluation of the tumor and have been shown to improve disease detection. Recent studies have demonstrated that plasma extracellular vesicle microRNAs (miRNAs) are functionally linked to cancer progression, metastasis, and aggressiveness. In our study, we analyzed a matched cohort with baseline blood plasma and MRI to access tumor morphology using imaging-based radiomics and cellular characteristics using miRNAs-based transcriptomics. Our findings indicate that the univariate feature-based model with the highest Youden’s index achieved average areas under the receiver operating characteristic curve (AUC) of 0.76, 0.82, and 0.84 for miRNA, MR-T2W, and MR-ADC features, respectively, in identifying clinically aggressive (Gleason grade) disease. The multivariable feature-based model demonstrated an average AUC of 0.88 and 0.95 using combinations of miRNA markers with imaging features in MR-ADC and MR-T2W, respectively. Our study demonstrates combining miRNA markers with MRI-based radiomics improves predictability of clinically aggressive prostate cancer.

## INTRODUCTION

Prostate cancer (PCa) is the second most common cause of cancer mortality among men worldwide and represents a significant health burden^1^. PCa is a heterogeneous disease that can manifest as either a low-risk, indolent tumor; about 42-66% of patients are estimated to be indolent Pca^2^ or as a high-risk, aggressive tumor that may eventually metastasize and become lethal if untreated. The widespread adoption of serum-based prostate-specific antigen (PSA) tests has significantly improved early detection of PCa^3^. However, the PSA test lacks specificity, which has led to a higher rate of false detection of tumors^4, 5^. Most localized PCa patients with higher Gleason grade tend to prophetically obtain radical prostatectomy (RP) as a curative option. In some of these patients, the disease progresses to present as biochemical recurrence (BCR) with an increased risk of metastasis^6–9^. It becomes critical to have reliable biomarkers capable of diagnosing clinically significant diseases early and can distinguish disease progression, which will greatly improve patient outcomes^10^.

Due to its ability to assess the whole prostate gland, magnetic resonance imaging (MRI) has been adopted as the primary modality to clinically stage prostate disease^11^. The prostate imaging reporting and data system (PIRADS) allows radiological assessment of prostate disease, which has improved standardized reporting but still suffers from inter-reader variability^12^. Radiomics has evolved as a methodology to characterize tumor morphology, and these quantitative metrics can prognosticate disease progression in oncological diseases^13–15^.

Extracellular vascular microRNAs (miRNAs) are short, non-coding RNA molecules that regulate gene expression post-transcriptionally^16^. They play critical roles in various cellular processes and are involved in the pathogenesis of numerous diseases, including cancer^16, 17^. miRNAs are typically found inside cells. However, some are shed into circulation in lipid-coated particles known as exosomes^18^. Circulatory exosomal miRNAs have been identified as possible disease biomarkers as they are relatively stable in blood and are protected from endogenous RNase activity. Recently, several miRNAs have been implicated as crucial regulators in PCa progression, some targeting oncogenes with an impact on cancer proliferation^19–22^. These miRNAs have been shown to target the most common oncogene pathways like the mTOR pathway^23^ and cell cycle regulation^24, 25^. These findings underscore the diverse roles of miRNAs in PCa pathogenesis and therapeutic responses.

In this study, we obtained radiomic characterization of abnormal regions using baseline biparmetric MRI. We also quantified the exosomal miRNAs in blood plasma in the same cohort of patients. In combination, we showed that these non-invasive complementary assessments (imaging, miRNAs) can predict clinically significant prostate disease at the patient level. We also outline the role of multi-omic features using biparametric MRI (MR-T2w, MR-ADC) features and blood plasma (miRNAs) to provide improved predictability and allow better reproducibility across patients.

## MATERIAL AND METHODS

### Patient Cohort and Plasma Sample Preparation

The patient cohort for the study was retrospectively obtained from The Moffitt Cancer Center. Patients were enrolled in the institutional research protocol (Total Cancer Care), which waived additional informed consent for the research study. Our retrospective research protocol allows access to the presented study, approved by the Moffitt Cancer Center/University of South Florida’s Institutional Review Board (IRB). Diagnostic multiparametric Magnetic resonance imaging (MRI) was obtained before treatment or biopsy. We selected patients who had prostate MRIs for this study to get the best characterization of the gland region. The patients clinical record and pathological assessment of the biopsy specimen were obtained from the medical record.

Blood samples (5-10 mL) were collected in EDTA K2 vacutainers from patients diagnosed with PCa (n=48), prior to treatment. Plasma samples were processed by initial centrifugation at 1300× g for 10 minutes at room temperature (RT). The resulting plasma was then transferred to a fresh 1.5 mL centrifuge tube and subjected to a second centrifugation step at 5000× g for 10 minutes at RT to obtain platelet-poor plasma. Aliquots of 250 µL from the processed plasma were quickly preserved at −80°C until subsequent processing for exosome isolation. We formed patient sub-cohorts with following groups: MR imaging, blood/plasma, matched imaging and blood plasma.

### Prostate Lesion delineation

The multiparametric MR imaging was assessed by our clinical radiologists (JC and TG), who provided consensus reading for the most aggressive (pathological grade) disease regions, along with glandular boundaries on the prostate MRI (T2W). The annotations were digitally recorded as an RT (radiotherapy format) referenced to T2W and stored on our research PACS (MIM software®). The clinical reports (radiology, pathology) were available for the clinical radiological assessment. We used bi-parametric MR modalities for the study, MR-ADC modality was semi-automatically registered to T2W using intensity-based image registration in Matlab® and the image resolution was remapped to T2W, used as a reference.

### Prostate Imaging and Quantification

Baseline bi-parametric MRI on patients with blood specimens were collected before treatment for PCa. The imaging cohort was restricted to the patients who followed the prostate MR image protocol to have a better characterization of prostate glandular anatomy. Our institutional clinical radiologists (JC, TK) reviewed the patient’s imaging (T2W, diffusion-weighted imaging or DWI/ Apparent diffusion coefficient or ADC) and identified the abnormalities across the prostate glandular anatomy. The abnormal regions were digitally recorded spans across the glandular volume (3 Dimensions) in RT (radiation therapy) format and stored on our research Picture Archive Communication System (PACS) (MIM Software Inc.). Institutional in-house radiomics feature extraction (306 features) was used to quantify abnormal regions of interest on the bi-parametric MR imaging (T2, ADC), which complies with the recommendations of the Image Biomarkers Standardization Initiative (IBSI)^26^. The radiomics toolbox has 306 quantitative features spanning three major categories (size, shape and texture), that was independently extracted in each of the biMRI modalities (MR-T2W, MR-ADC).

### Plasma Exosome Isolation

The exosome isolation process began with the thawing of 250 µl plasma aliquots at room temperature (RT), followed by their transfer to 1.5 mL microcentrifuge tubes. These tubes were then centrifugated at 10,000× g for 15 minutes at 4°C to eliminate large vesicles and cellular debris, yielding a supernatant utilized for exosome isolation. The SBI SmartSEC^TM^ Single for EV Isolation^TM^ (System Biosciences, Palo Alto, CA, USA; cat# SSEC200A-1) was employed as a size exclusion chromatography-based approach. The exosomes were eluted using phosphate buffer saline (PBS) and stored at −-80°C until miRNA extraction. The plasma samples were randomized before processing, which will mitigate batch effects in our analysis.

### miRNA Extraction

The extraction of exosome miRNA was conducted using the miRNeasy (Micro) Kit (Qiagen, Valencia, CA, USA, cat# 217084). Initially, 200 µL of the exosome sample was mixed with 1 mL of QIAzol Lysis Reagent, followed by chloroform addition and centrifugation at 12,000 × g for 15 minutes at 4°C to isolate the RNA-containing aqueous phase. The extracted RNA underwent purification using the RNeasy Mini Elute spin column, involving ethanol washes and specific buffers, and elution with 15 µl of RNase-free water. The concentration of RNA was measured using the QuantiFluor® RNA System (Promega, Madison, WI, USA, Cat#E3310) with Quantus equipment, and the eluted RNA was subsequently stored at −80°C.

### Library preparation

miRNA libraries were generated employing the QIAseq miRNA library kit (Qiagen, Valencia, CA, USA cat#331502) with 5 µl of total RNA utilized for library preparation. The process involved initial ligation of 3’ and 5’ adapters to the miRNAs. Complementary DNA (cDNA) libraries were then constructed via reverse transcription, followed by 22 cycles of PCR amplification and subsequent cleanup of cDNA using QMN beads. The concentration of the prepared libraries was quantified using the Qubit 2.0 Fluorometer with the Qubit™ dsDNA Quantification Assay Kits (Thermo Fisher Scientific, Middletown, VA, USA, cat#Q32851). Additionally, library quality was assessed using the Agilent High Sensitivity DS1000 method and the Agilent 2200 TapeStation (Agilent, Santa Clara, CA, USA, cat#5067-5585). Subsequently, libraries were pooled in an equimolar ratio based on their molarity, and the weight-to-moles conversion ratio for nucleic acids was determined.

### miRNA-seq and Data Analysis

The miRNA-seq procedure began with pooling 20 to 24 libraries, following the guidelines outlined in the NextSeq System - Denature and Dilute Libraries Guide. To ensure quality control, 1% PhiX Control v3 was incorporated into all pools as an internal standard. Single-read sequencing was performed with a 75 bp read length using the NextSeq 500 Sequencing System and the NextSeq 500/550 High Output v2.5 kit (75 cycles) (Illumina, San Diego, CA, USA cat#20024906).

Prior to alignment, the sequencing data’s quality control (QC) was executed using FastQC (version 0.11.9). Subsequently, adaptor removal was carried out using cutadapt (version 3.3). The adapter-trimmed small RNA sequencing reads were then mapped against the miRBase database (version 21) utilizing the DNAStar tool (version 3.2). All statistical analyses were conducted within the R environment (version R4.0.3).

### Biological Pathway Enrichment related to miRNAs

Regulatory targets and functional annotations of microRNAs were identified using TargetScan ^27^ and miRDB ^28^. The Database for Annotation Visualization, and Integrated Discovery (DAVID V 6.7) was used to identify functional biological pathways for top miRNAs identified by our analysis. Furthermore, miRanda software was utilized for target prediction of the putative novel microRNA sequences ^29, 30^.

### Redundancy reduction and Statistical methods

Coefficient of discrimination (R^2^) between the features was computed to quantify dependency across the patient samples in our cohort. the metric (R^2^) was iteratively computed between all possible features and highly dependent features (R^2^ ≥ 0.99) were flagged. In this dependent group, a representative feature with the highest variability across the patient population was selected, and others were removed. This process was repeated across each sub-cohort and modalities (miRNA, MR-T2W, MR-ADC)^31^. The process allowed forming a feature set that was uncorrelated (see Table 1). The level of dependency threshold needs to be balanced between removing correlated features and leaving behind those with information.

**Table 1.**
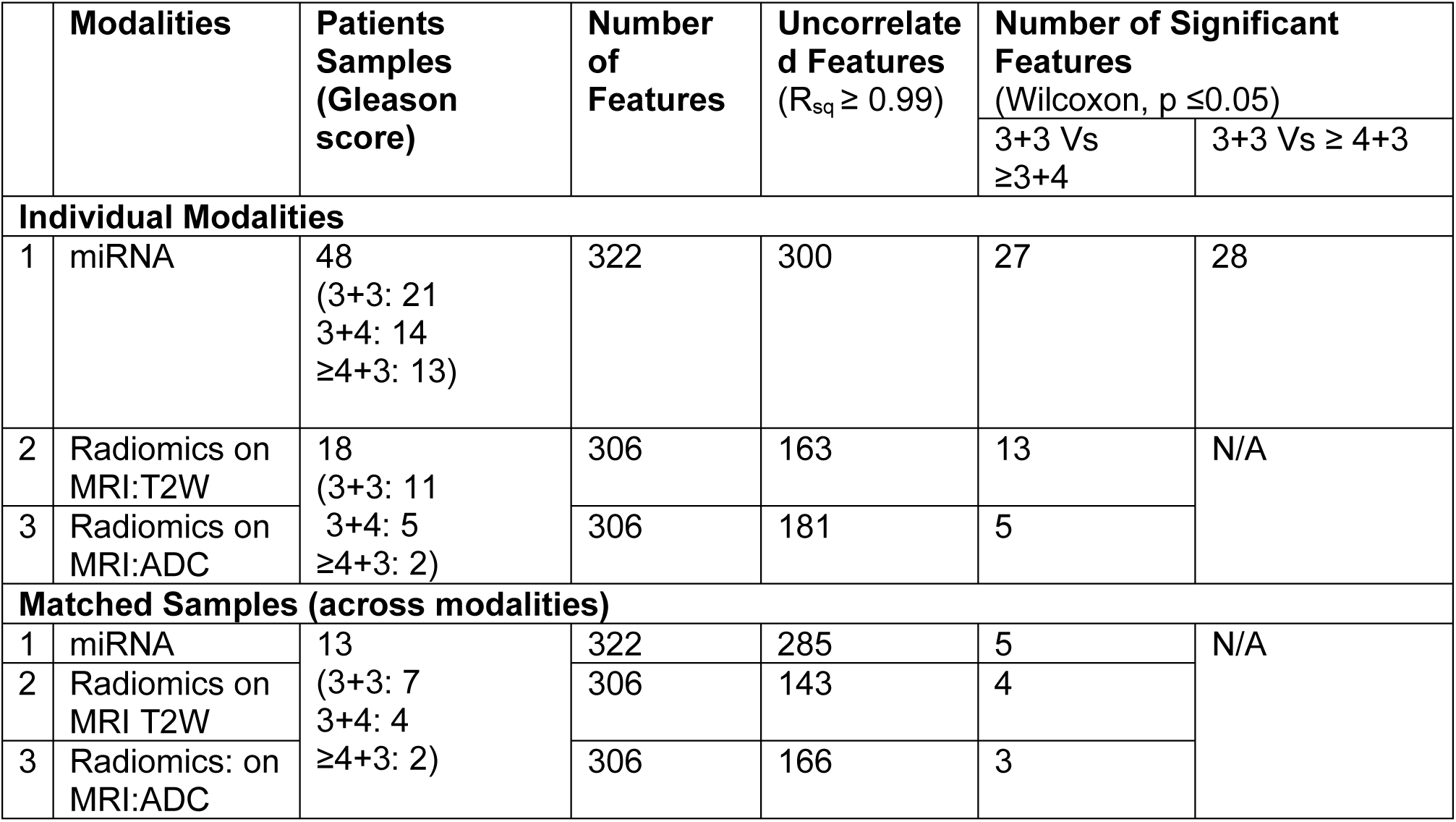
Feature that are non-dependent across fluid and imaging modalities.

a logistic regression-based classifier model in univariate and multivariable (up to three dimensions) was then built using uncorrelated features identified in our cohorts. All possible combinations of features were evaluated to find the best feature combination in each cohort, and this was repeated independently across the modalities. In our study, over 4.45 million possible pairs in the miRNA’s cohort, over 708 thousand pairs in the MR-T2W, and 971 thousand in the MR-ADC cohort were evaluated, respectively. The feature pair was sorted based on hold-out (80/20, train/test) test classification accuracy, and estimates were randomly repeated (over 200 times). Combination mixed multimodal features were then formed by selecting the top candidates from each combination (1, 2, and 3 pairs). Sensitivity, specificity, positive predictive value, negative predictive value, and area under receiver operator characteristics were estimated using cross-validation method with average estimates reported. The feature-based models were ranked based on Youden’s index (Sensitivity + Specificity −1) and receiver operator characteristics area under the curve (ROC AUC or AUC) ^32^. A hold-out cross validation approach (80% train, 20% test) was used to estimate the model performance, which was averaged over multiple repeats (over 200), and ensemble test statistics reported.

## RESULTS

### 1. Patient characteristics

The study included 48 primary PCa patients with pretreatment blood plasma samples and MR imaging using mixed protocol (pelvic, prostate, abdomen). Of the samples, we converged on 13 patients (18 biopsies) who had prostate MR imaging that followed standardized prostate imaging protocol (see **Table 1**). We assessed patients imaging in each of the bi-parametric modalities (miRNA, MRI-T2/ADC) that were matched with plasma-based markers to create subcohorts (miRNA with MR-T2W and miRNA with MR-ADC). We carried out statistical analysis to identify features that discriminate clinically significant PCas defined by Gleason scores (GS≥ 3+4) across these subgroups, considering them independently.

### 2. Modality base classifiers

To identify individual miRNAs and image features that were associated with aggressive PCa, we first performed correlation analysis and removed correlated features (R^2^>=0.99) across all possible features in a modality (miRNA, n=48; MR-T2W/ADC, n=18 biopsies). This step removed 6.8%, 46.7%, and 40.8% of the metrics, leaving us with 300, 163, and 181 uncorrelated features for miRNA, MR-T2W, and MR-ADC, modalities respectively. While in the matched cohort (imaging & miRNA, n=13), we had 285 (removed 11.4%), 143 (removed 53.2%), and 166 (removed 40.8%) uncorrelated features for miRNA, MR-T2W, MR-ADC modalities, respectively. We then performed non-parametric test and identified individual features that were statistically significant across indolent and clinically significant patients (Table 1 and Figure 1). We then built classifier models using logistic regression with univariate and multi-variable (2 and 3) features. Predictive ability of these models was assessed based on area under the receiver operator characteristics (AUC) using a cross-validation (hold out) approach. For univariate feature-based model using either miRNA or imaging modalities, we found that miRNAs (R193: miR-151a-5p, R46: miR-93-5p) based model had an average AUC in the range of 0.66-0.76. MR-T2W radiomic features (Laws-features) had an average AUC range from 0.78 to 0.87, while MR-ADC radiomic features (Co-Occurrence, volume, wavelet) showed an average AUC range from 0.78 to 0.84 (see Table 2). An example univariate feature-based classifiers are shown in Figure 3.

**Figure 1.**
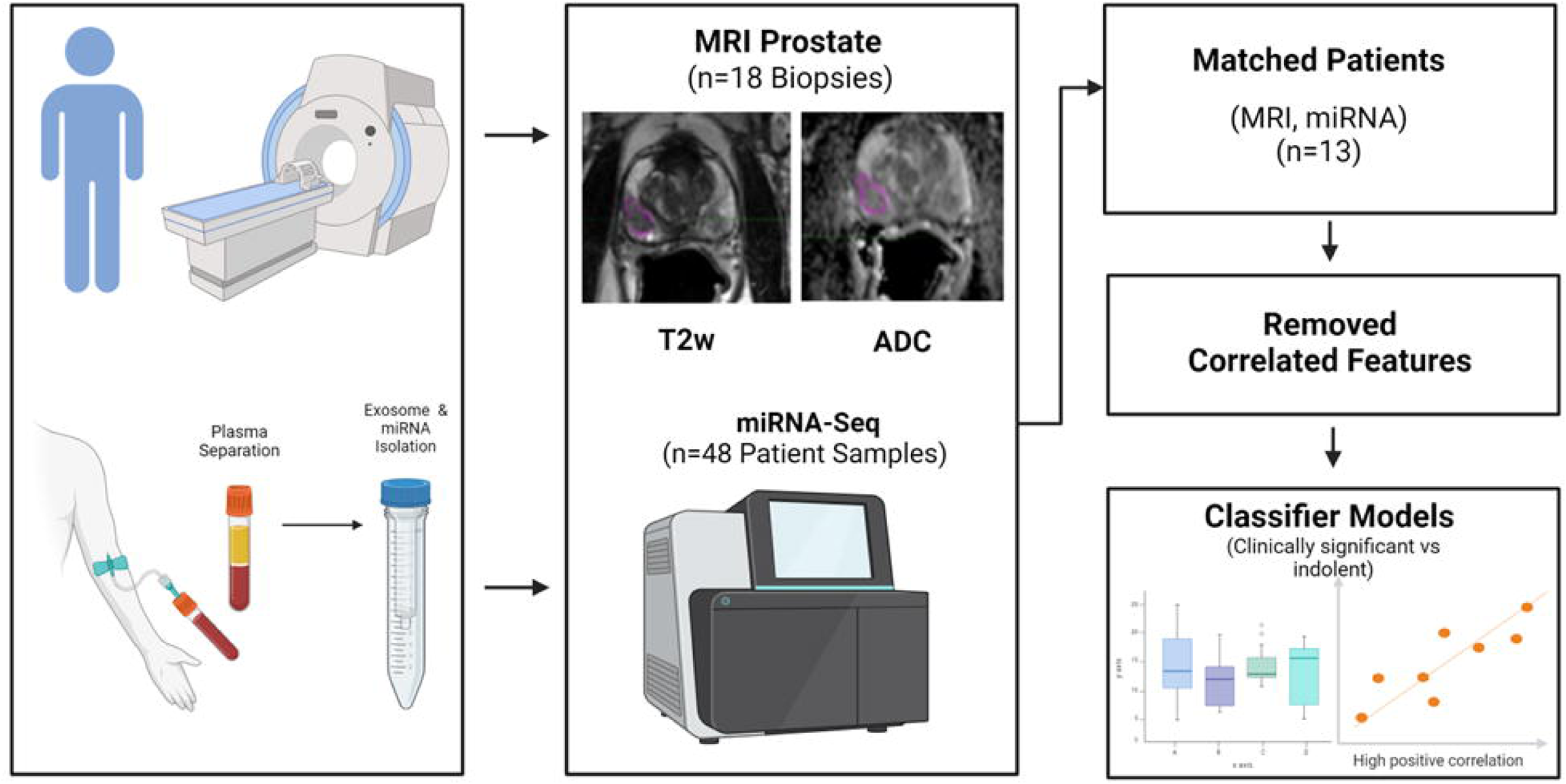
Process flow to identify discriminate features from multimodalities, plasma-based miRNA and MR image based radiomics (T2W, ADC) to discriminate aggressive grade prostate disease.

**Figure 2.**
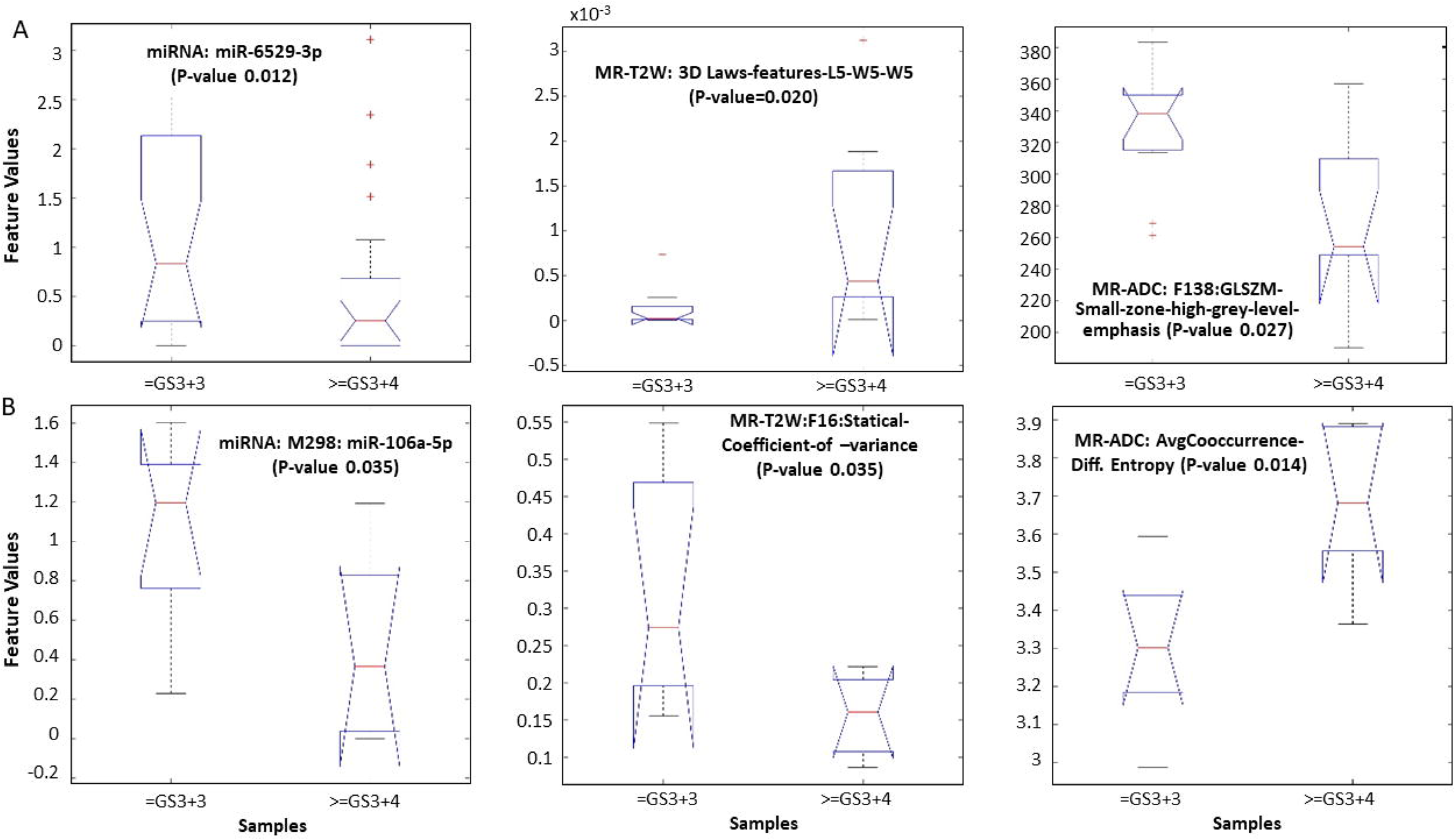
Feature distribution that differentiates clinically significant (>=3+4) from indolent (3+3) for a) individual cohorts (miRNA, MR T2w, MR ADC), b) combined cohort (miRNA, MR T2w, MR ADC).

**Table 2.** Features that discriminate clinically significant prostate cancer from indolent (3+3) using independent features in these modalities; a) miRNA, in 3+3 Vs ≥3+4 (n=48), b) miRNA, in 3+3 Vs ≥4+3 (n=34), c) MRI T2 W radiomics (n=18) d) MRI ADC radiomics (n=18).

| **(a) miRNA – Univariate (n=48) (GS 3+3 Vs $\geq 3+4$)** |  |  |  |  |
| --- | --- | --- | --- | --- |
|  | **miRNA (1-marker)** | **E[Sensitivity]/**<br>**E[Specificity]** | **E[PPV]/**<br>**E[NPV]** | **E[AUC], CI , Std** |
| 1 | miR-151a-5p | 0.798/0.68 | 0.756/0.726 | 0.76[0.388,0.956],0.162 |
| 2 | miR-338-3p | 0.807/0.573 | 0.696/0.709 | 0.699[0.213,0.956],0.201 |
| 3 | miR-93-5p | 0.775/0.547 | 0.692/0.67 | 0.767[0.457,0.971],0.15 |
| 4 | miR-208a-5p | 0.833/0.482 | 0.671/0.686 | 0.658[0.2,0.95],0.194 |
| 5 | miR-190a-5p | 0.842/0.469 | 0.654/0.687 | 0.702[0.392,0.968],0.173 |
| **(b) miRNA – Univariate (n=34) (GS 3+3 Vs $\geq 4+3$)** |  |  |  |  |
|  | **miRNA (1-marker)** | **E[Sensitivity]/**<br>**E[Specificity]** | **E[PPV]/**<br>**E[NPV]** | **E[AUC], CI , Std** |
| 1 | miR-93-5p | 0.684/0.857 | 0.762/0.798 | 0.821[0.3,0.95],0.208 |
| 2 | miR-8072 | 0.631/0.855 | 0.74/0.767 | 0.776[0.05,0.95],0.237 |
| 3 | miR-328-3p | 0.589/0.861 | 0.727/0.73 | 0.798[0.417,0.958],0.178 |
| 4 | miR-1469 | 0.574/0.845 | 0.652/0.772 | 0.772[0.333,0.958],0.183 |
| 5 | miR-5583-5p | 0.539/0.821 | 0.645/0.728 | 0.786[0.25,0.958],0.202 |
| **c) Univariate Imaging T2w (n=18) (GS 3+3 Vs $\geq 3+4$)** |  |  |  |  |
|  | **T2 (Radiomics)** | **E[Sensitivity]/**<br>**E[Specificity]** | **E[PPV]/**<br>**E[NPV]** | **E[AUC], CI , Std** |
| 1 | 3D_Laws_features_L5_E5_S5 | 0.588/0.882 | 0.627/0.748 | 0.83[0.05,0.95],0.267 |
| 2 | 3D_Laws_features_L5_E5_W5 | 0.522/0.897 | 0.645/0.713 | 0.871[0.325,0.963],0.192 |
| 3 | 3D_Laws_features_L5_S5_S5 | 0.575/0.802 | 0.587/0.712 | 0.827[0.05,0.95],0.264 |
| 4 | 3D_Laws_features_L5_W5_W5 | 0.432/0.925 | 0.565/0.688 | 0.853[0.2,0.95],0.238 |
| 5 | 3D_Laws_features_E5_E5_L5 | 0.448/0.892 | 0.557/0.699 | 0.778[0.05,0.95],0.312 |
| **d) Univariate Imaging ADC (n=18) (GS 3+3 Vs ≥3+4)** |  |  |  |  |
|  | **ADC (Radiomics)** | **E[Sensitivity]/**<br>**E[Specificity]** | **E[PPV]/**<br>**E[NPV]** | **E[AUC], CI , Std** |
| 1 | Volume_density_minimum_volume_enclosing_ellipsoid | 0.72/0.832 | 0.702/0.808 | 0.844[0.367,0.967],0.208 |
| 2 | Maximum_histogram_gradient_grey_level | 0.413/1 | 0.61/0.708 | 0.777[0.525,0.975],0.174 |
| 3 | GLSZM_Small_zone_high_grey_level_emphasis | 0.582/0.81 | 0.663/0.693 | 0.818[0.3,0.95],0.234 |
| 4 | avgCooccurrence_Difference_entropy | 0.605/0.758 | 0.546/0.759 | 0.801[0.05,0.95],0.263 |
| 5 | 3D_Wavelet_P1_L2_C12 | 0.648/0.71 | 0.576/0.714 | 0.817[0.325,0.963],0.22 |

**Figure 3.**
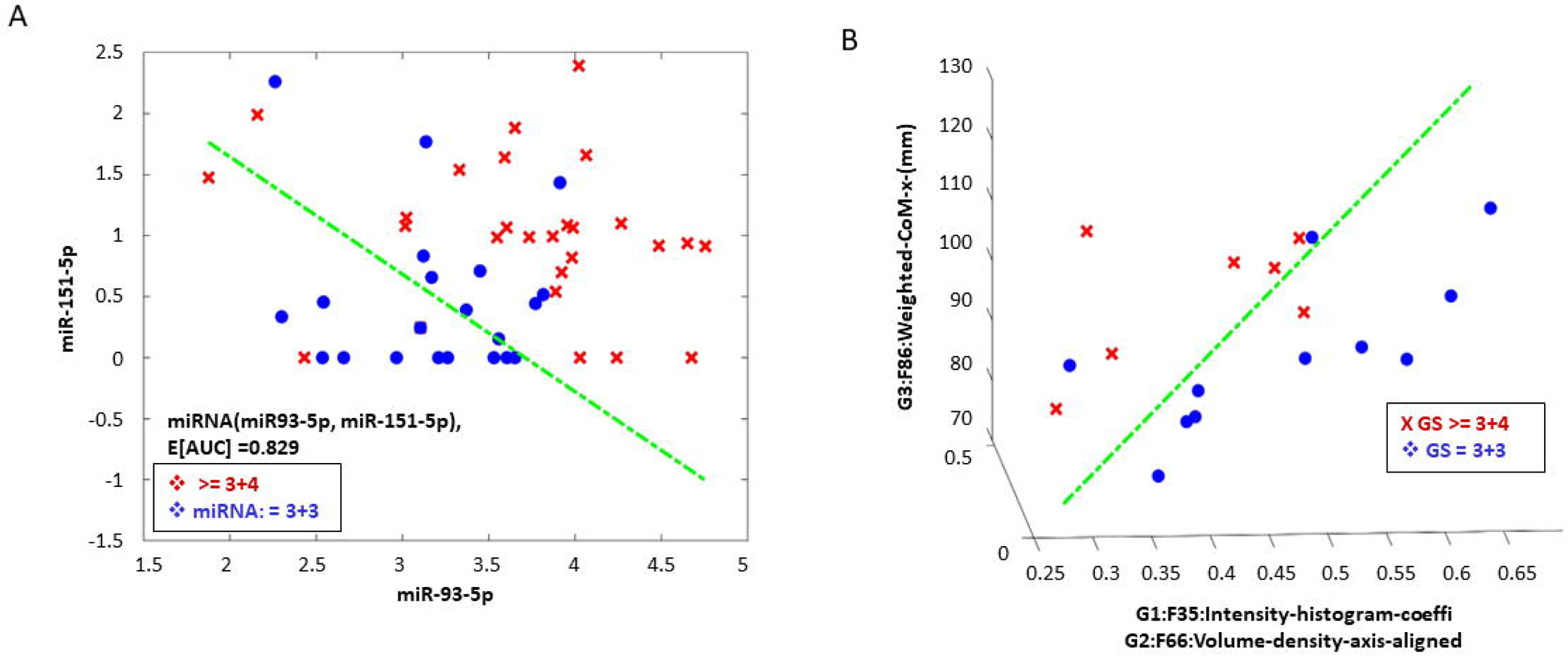
Multi-feature scatter plot to show spread of aggressive from indolent grade prostate cancer along with discrimination boundary A) using miRNA-based features (miR-93-5p, miR-151a-5p) b) MR-T2w features (3-features).

### 3. Multimodal classifier model

To evaluate if combination of multi-modality features could improve performance of detecting aggressive disease, we built multi-modal predictors by combining miRNAs and bi-parametric MR features (MR-T2W / ADC) from respective modalities in the matched cohort (n=13). In multimodal feature analysis, we selected the best univariate miRNA’s that had functional relevance to prostate oncology. Using miRNA and MR-T2W radiomics, univariate features had an average AUC of 0.65 to 0.71 and 0.77 to 0.90, respectively. Combination of these two features (miRNA & MR-T2W radiomics) had an average AUC of 0.73 to 0.86. Using two feature-based models (miRNA: miR-7704, miR-151a-5p, T2W: COV, Co-Occurrence) from each of the modalities, seems to moderately complement AUC (average range from 0.79 to 0.96). While using a single feature from miRNA and MR-ADC modalities, a combination (miRNA & MR-ADC) had an average AUC range from 0.73 to 0.75, 0.87 to 0.90 and 0.77 to 0.88, respectively. While using two features from each of the modalities (miRNA: miR-151a-5p, miR-338-3p & MR-ADC: Co-occurrence, Laws features) improved AUC (average range from 0.76 to 0.88) in comparison on using them individually. Importantly, the sensitivity/specificity was higher compared to individual modality-based models (see Table 3 and Figure 4).

**Table 3.** Multimodal features based (miRNA and imaging) model to discriminate clinically significant prostate cancer (≥3+4) from indolent (3+3) in a matched cohort of patients (n=13). (A1&2) extracellular exosomal miRNA with MRI T2W. (B-1&2) extracellular exosomal miRNA with MRI ADC features.

| **(A1) Combined: miRNA + Img (T2W): (GS 3+3 Vs $\geq 3+4$): 1-dimension** |  |  |  |  |  |
| --- | --- | --- | --- | --- | --- |
|  | miRNA | Imaging (T2W) | E[AUC], CI , Std |  |  |
|  |  |  | miRNA | Img: T2W | Combined (miRNA, T2W) |
| 1 | miR-151a-5p | avgCooccurrence_Joint_MAX | 0.713[0.05,0.95],0.347 | 0.77[0.05,0.95],0.358 | 0.73[0.05,0.95],0.344 |
| 2 | miR-7704 | Statistical_Coefficient_of_variance | 0.653[0.05,0.95],0.371 | 0.9[0.4,0.95],0.225 | 0.858[0.4,0.95],0.223 |
| **(A2) Combined: miRNA + Img (T2W): (GS 3+3 Vs $\geq 3+4$): 2-dimension** |  |  |  |  |  |
|  | miRNA | Imaging (T2W) | E[AUC], CI , Std |  |  |
|  |  |  | miRNA | Img: T2W | Combined (miRNA, T2W) |
| 1 | miR-151a-5p;M275:miR-6717-5p; | Volume_at_intensity_fraction_10;F86:Weighted_CoM_x_(mm); | 0.978[0.7,0.95],0.118 | 0.948[0.525,0.975],0.134 | 0.95[0.2,0.95],0.178 |
| 2 | miR-7704;M189:miR-3136-3p; | avgCooccurrence_Autocorrelation;F301:3D_Wavelet_P1_L2_C12; | 0.633[0.05,0.95],0.368 | 0.963[0.525,0.975],0.12 | 0.958[0.525,0.975],0.128 |
| 3 | miR-151a-5p;M252:miR-338-3p; | GLSZM_High_grey_level_zone_emphasis;F301:3D_Wavelet_P1_L2_C12; | 0.573[0.05,0.95],0.406 | 0.898[0.05,0.95],0.219 | 0.785[0.4,0.95],0.238 |
| **(B1) Combined: miRNA + Img (ADC): (GS 3+3 Vs $\geq 3+4$): 1-dimension** |  |  |  |  |  |
|  | miRNA | Imaging (ADC) | E[AUC], CI , Std |  |  |

|  |  |  | miRNA | Img: ADC | Combined (miRNA, ADC) |
| --- | --- | --- | --- | --- | --- |
| 1 | miR-338-3p | avgCooccurrence_Difference_entropy | 0.75[0.05, 0.95], 0.314 | 0.865[0.05, 0.95], 0.29 | 0.88[0.475, 0.963], 0.212 |
| 2 | miR-151a-5p | Volume_density_minimum_volume_enclosing_ellipsoid | 0.733[0.05, 0.95], 0.318 | 0.895[0.2, 0.95], 0.241 | 0.768[0.05, 0.95], 0.312 |
| **(B2) Combined: miRNA + Img (ADC): (GS 3+3 Vs ≥3+4): 2-dimension** |  |  |  |  |  |
|  | miRNA (2-dim) | Imaging (ADC) – (2-dim) | E[AUC], CI , Std |  |  |
|  |  |  | miRNA | Img: ADC | Combined (miRNA, ADC) |
| 1 | miR-190b-5p; | avgCooccurrence_Difference_entropy; | 0.843[0.05, 0.95], 0.272 | 0.88[0.05, 0.95], 0.276 | 0.88[0.4, 0.95], 0.237 |
|  | miR-106b-3p | avg_3D_LGRE_(Low_grey_level_run_emphasis) |  |  |  |
| 2 | miR-151a-5p; | 3D_Laws_features_E5_E5_W5; | 0.6[0.05, 0.95], 0.381 | 0.898[0.05, 0.95], 0.256 | 0.76[0.2, 0.95], 0.261 |
|  | miR-338-3p | 3D_Laws_features_E5_R5_E5 |  |  |  |

**Figure 4.**
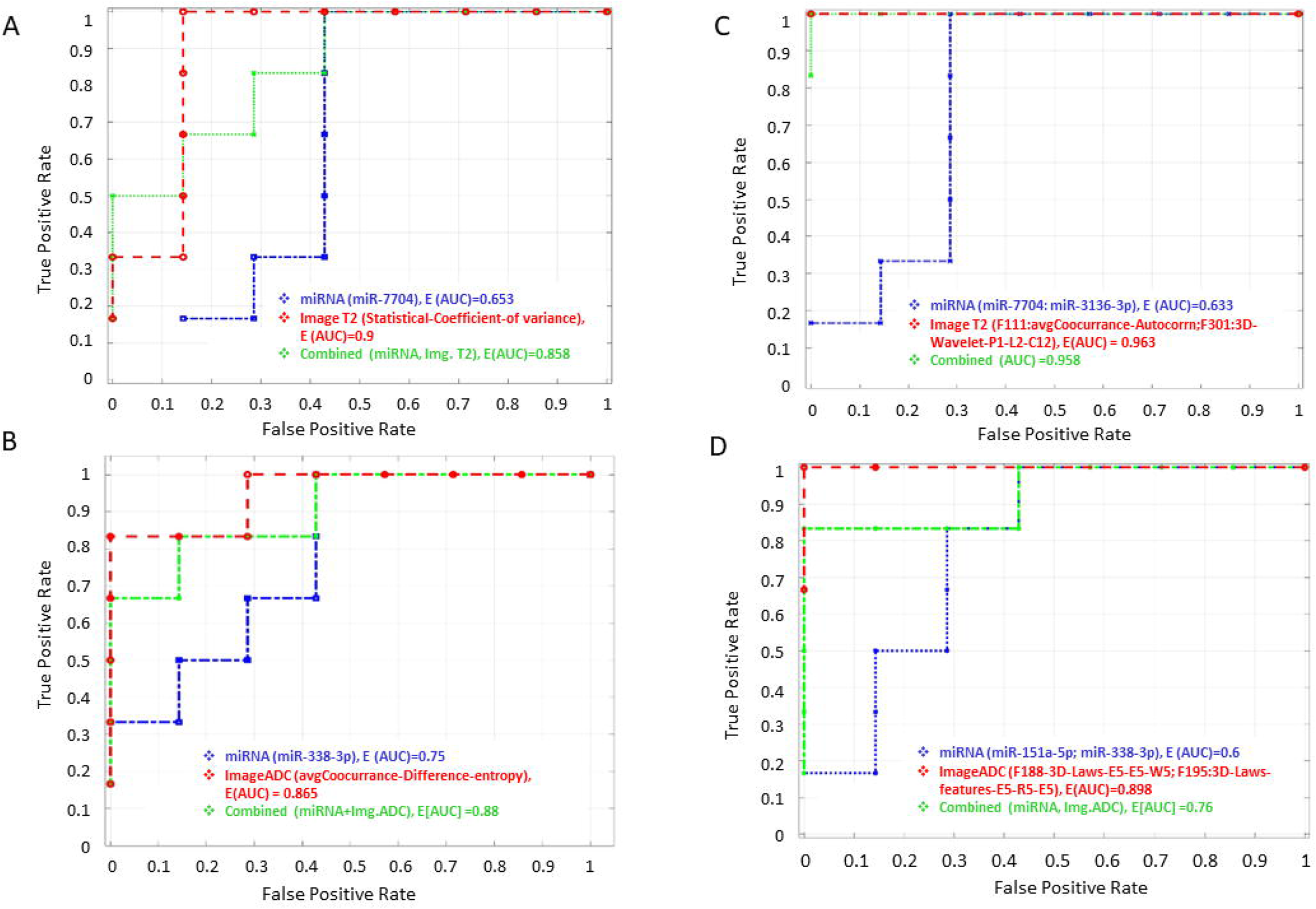
Receiver operating characteristic curve (ROC) in using top predictors to discriminate clinically significant (Gleason 3+4) from indolent grade prostate cancers A) miRNA (hsa-miR-7704) with T2W radiomics (univariate), C) miRNA (has-miR-338-3p) with ADC radiomics (univariate).

### 4. Gene Ontology and regulatory pathways

After identifying top miRNAs (miR-151a-5p, miR-338-3p, miR-7704, miR-93-5p, and miR-190b-5p) that were predictors of aggressive PCa we used these markers to link regulatory pathways associated with their predicted targets using the following curated databases: TargetScan (targetscan.org) and miRDB target computational prediction software. We also evaluated other open-source pathway miner tools (KEGG^33^, PANTHER^34^, and Database for Annotation, Visualization, and Integrated Discovery, DAVID^35^). The data-mining analysis identified the most relevant pathways using miRNAs as seeds that were most common between TargetScan and miRDB. We found a significant enrichment in the following gene-pathway associations: Pathways in cancer, PI3K, Akt signaling, FoxO signaling and Wnt signaling pathway genes, reported by PANTHER (See Figure 5A). In addition, ras signaling, angiogenesis, FGF signaling, wnt signaling, and PDGF (Platelet-derived growth factor) signaling pathway were the most significant pathways obtained using the KEGG pathway (see Figure 5B).

**Figure 5.**
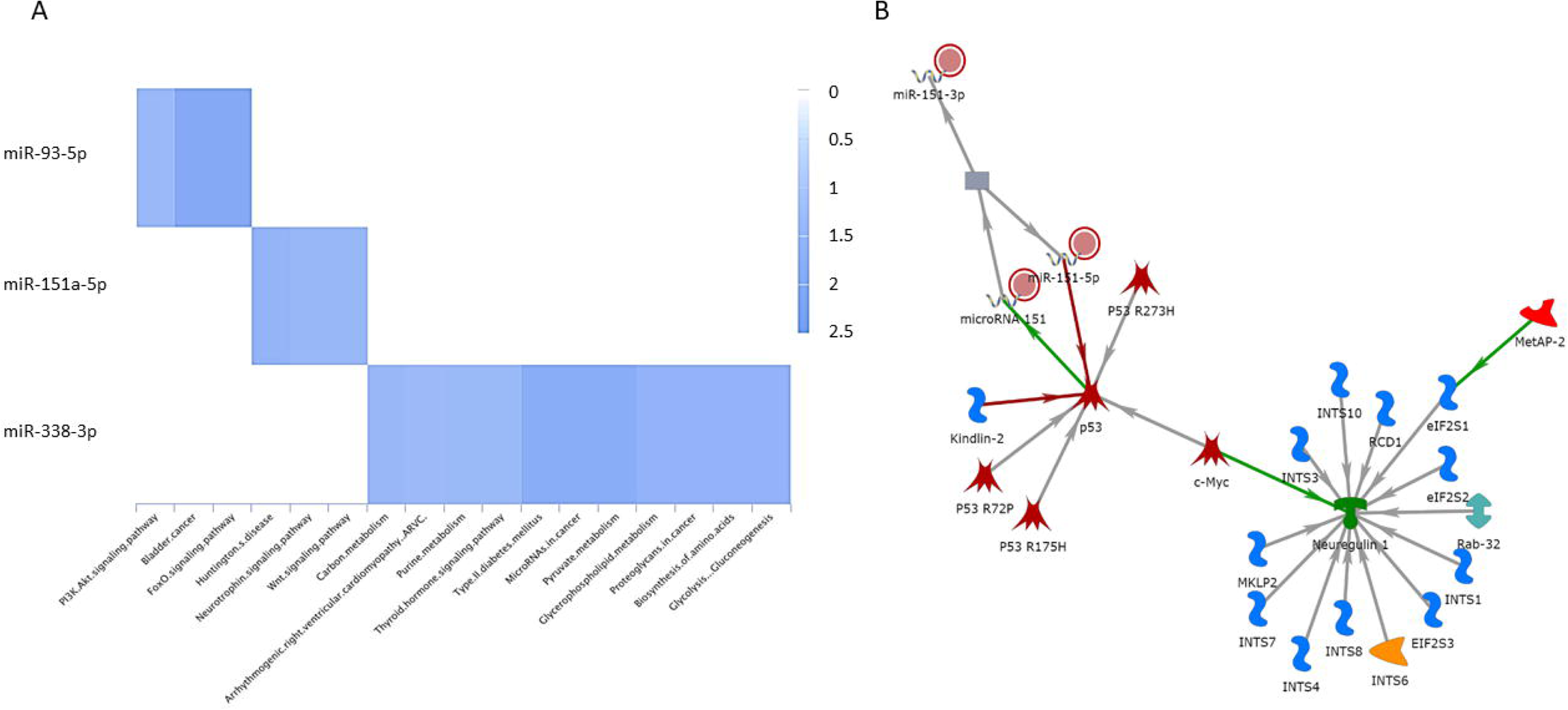
Regulators pathways enriched for miRNAs (miR-151a-5p, miR-338-3p, miR-7704, miR-93-5p, miR-190b-5p), that was identified in our predictive analysis. Pathway tools using; a) GO functional pathway (Pather.org), b) p52 pathway enriched, seeded with two miRNAs (miR-151a-5p, miR-338-3p) using (GeneGo®).

## DISCUSSION

PCa diagnosis and treatment strategies have been improved in the last two decades^36–39^. Despite these changes, early detection of clinically significant cancers remains challenging^39^.Since the use of PSA-based tests has resulted in a high level of false diagnosis^3, 40^,genomic-based technologies are believed to provide a promising tool in identifying aggressive disease^37, 41^. Clinical use of genomic markers to assess metastasis of the disease has improved the management of the disease^42^. For example, the use of extracellular miRNAs has evolved in the assay development for disease detection, including PCa^25, 43^. Prior studies have shown radiomic features related to histogram intensity and cooccurrences were predictive of aggressive prostate disease, and these metrics have been related to biochemical recurrence^44, 45^. Recent work has implicated radiomic metrics related to first-order statistics, texture (laws features, Haralick/cooccurrence) features extracted in MR-T2w, and radiomics features related to texture, edge descriptors (Laws, gradient, Sobel) computed in MR-ADC were associated to aggressive disease grades^45^. In comparison, our study finds several imaging features related to intensity and texture-based features (co-occurrence, wavelets, Laws) in T2W/ADC modalities were predictive of aggressive prostate disease (see **Tables 2 & 3**).

Numerous studies have demonstrated that specific miRNAs are differentially expressed in PCa, making them valuable for early diagnosis and disease monitoring. Our study, utilizing a univariate feature-based model with miRNA and imaging modalities, identified miR-151a-5p and miR-93-5p as having the highest AUC (see **Table 2**). Other possible combinations of miRNA and imaging features are deferred to supplemental section (See Supp. Table ST.1). miR-151a-5p is differentially expressed in PCa, indicating its role in tumor aggressiveness^46^. This miRNA is well-known as an oncogene, particularly in colorectal cancer, and is also overexpressed in lung cancer and lymphoblastic leukemia^47–49^. Our KEGG analysis predicted that miR-151a-5p indirectly targets the Neuregulin 1 (NRG1) gene through P53 and c-Myc. Recent studies have also shown that the NRG1 gene promotes antiandrogen resistance in PCa^50^. The consistent dysregulation of miR-151a-5p across various cancers highlights its universal role in oncogenic processes. The overexpression of miR-93-5p has been linked to increased migration and invasion in squamous cell carcinoma of the head and neck, suggesting an oncogenic role^51, 52^. While the exact mechanism of action needs further investigation, elevated levels of miR-93-5p have also been associated with epithelial-mesenchymal transition (EMT), radiotherapy response, and poor prognosis^52–54^. Additionally, miR-93-5p has been shown to be upregulated in oral cancer^55, 56^. These findings reinforce the role of miRNAs in regulating PCa, a foundation for further research into their mechanistic roles and therapeutic potential (See Table 4).

**Table 4.**
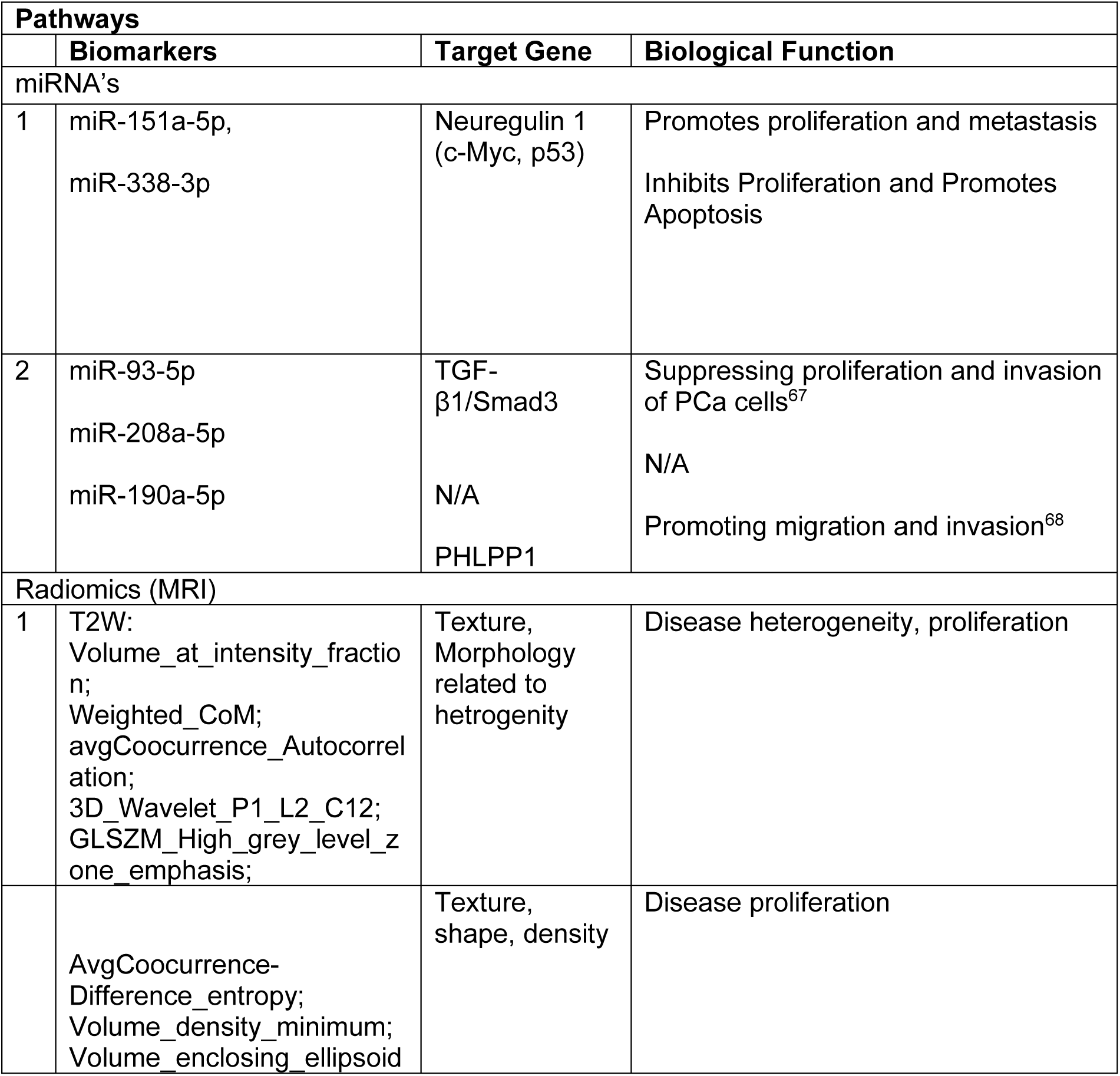
Predictive biomarkers relationship to biological pathways.

| Pathways |  |  |  |
| --- | --- | --- | --- |
|  | Biomarkers | Target Gene | Biological Function |
| miRNA's |  |  |  |
| 1 | miR-151a-5p,<br>miR-338-3p | Neuregulin 1<br>(c-Myc, p53) | Promotes proliferation and metastasis<br>Inhibits Proliferation and Promotes Apoptosis |
| 2 | miR-93-5p<br>miR-208a-5p<br>miR-190a-5p | TGF-β1/Smad3<br>N/A<br>PHLPP1 | Suppressing proliferation and invasion of PCa cells<sup>67</sup><br>N/A<br>Promoting migration and invasion<sup>68</sup> |
| Radiomics (MRI) |  |  |  |
| 1 | T2W:<br>Volume_at_intensity_fraction;<br>Weighted_CoM;<br>avgCooccurrence_Autocorrelation;<br>3D_Wavelet_P1_L2_C12;<br>GLSZM_High_grey_level_zone_emphasis; | Texture,<br>Morphology<br>related to<br>heterogeneity | Disease heterogeneity, proliferation |
|  | AvgCooccurrence-Difference_entropy;<br>Volume_density_minimum;<br>Volume_enclosing_ellipsoid | Texture,<br>shape, density | Disease proliferation |

In our multi-modal miRNA and bi-parametric MR feature-based analysis, we identified miR-7704, miR-3136-3p, miR-151a-5p, and miR-338-3p as having high AUC values. Based on other studies, miR-7704 emerges as a potential target for aggressive PCa, consistent with its reported roles in ovarian and breast cancers^57, 58^. In ovarian cancer, miR-7704 is part of a feedback loop with IL2RB and AKT, influencing tumorigenesis and chemoresistance^57^. This suggests that miR-7704 may play a critical role in cancer progression and underscores its potential as a therapeutic target and prognostic biomarker across different cancer types. In another study, miR-3136-3p was significantly upregulated in high-grade cervical intraepithelial neoplasia in liquid biopsy samples^59^. miR-338-3p is downregulated in several cancers, including gastric, ovarian, and breast cancers^60–63^. In PCa cells, overexpression of the miR-338-3p suppresses cell migration and invasion^64–66^. These miRNA with high AUC in multi-modal miRNA and bi-parametric MR feature-based analysis, indicating their strong diagnostic potential for aggressive prostate cancer (PCa). These miRNAs may serve as valuable therapeutic targets and prognostic biomarkers across various cancer types (see Table 3 & 4 and Figures 3&4).

Although single omics analysis has shown promising in identifying aggressive disease, the PCa is known to be highly heterogenous. One omics data may not capture the complete landscape of PCa biology (add some references). It is believed that multi-omic approach can increase sensitivity of biomarkers. Therefore, we evaluated the multi-omic approach in a matched cohort of pre-treatment blood plasma and MR imaging to identify biomarkers in localized PCa patients. Our data showed that several miRNA and image-based features are differentially expressed in clinically significant PCa. By combining circulating extracellular transcriptomes and MRI-based radiomes, our multi-omic model improved performance in distinguishing aggressive disease. The integration of these features significantly enhances the accuracy of predicting clinically significant PCa, demonstrating the value of a mixed-modality approach in assessing disease aggressiveness.

### Limitations

Our study has relatively small sample size, which may affect the generalizability of the findings. Additionally, the reliance on a single institutional cohort may introduce bias and limits the racial cross-sectional nature of the data that could add to biases. We used several mitigation strategies that include cross validation approach. The study also did not account for potential confounding factors such as providers, patient racial/treatment history and genetic variability. Further research with larger, diverse cohorts and longitudinal data is necessary to validate and expand upon these results.

## CONCLUSION

Our study findings highlight the significant roles of circulating transcriptomics and radiomics in identifying aggressive PCa. By outlining specific miRNAs and MR imaging features, the study enhances our understanding of PCa pathogenesis through improved assessment of morphological characteristics. The combination of prostate miRNAs with imaging metrics offers a non-invasive method for assessing aggressive disease. However, validation of these findings in secondary, independent studies is essential.

## Data Availability

All data produced in the present study are available upon reasonable request to the authors.

## ETHICS STATEMENT

This study was approved by the Moffitt Cancer Center IRB (protocol number mcc#18104).

## Authors Contributions

KKA, LW: miRNA assay and quantification

JC, TG: Radiological annotation and opinion

YB, JC: Imaging metrics and quantification

YB, KKA, LW: statistical analysis, inference and hypothesis

KKA, LW, YB, BJM, RG: manuscript writing/editing and approval.

## REFERENCES

1. Siegel RL, Miller KD, Wagle NS, Jemal A. Cancer statistics, 2023. CA Cancer J Clin. 2023;73(1):17–48. Epub 2023/01/13. doi: 10.3322/caac.21763. PubMed PMID: 36633525.

2. Stenzl A, Studer UE. Outcome of patients with untreated cancer of the prostate. Eur Urol. 1993;24(1):1–6. Epub 1993/01/01. doi: 10.1159/000474253. PubMed PMID: 8365427.

3. Ilic D, Djulbegovic M, Jung JH, Hwang EC, Zhou Q, Cleves A, Agoritsas T, Dahm P. Prostate cancer screening with prostate-specific antigen (PSA) test: a systematic review and meta-analysis. BMJ. 2018;362:k3519. Epub 20180905. doi: 10.1136/bmj.k3519. PubMed PMID: 30185521; PMCID: PMC6283370.

4. Draisma G, Etzioni R, Tsodikov A, Mariotto A, Wever E, Gulati R, Feuer E, de Koning H. Lead time and overdiagnosis in prostate-specific antigen screening: importance of methods and context. J Natl Cancer Inst. 2009;101(6):374–83. Epub 2009/03/12. doi: 10.1093/jnci/djp001. PubMed PMID: 19276453; PMCID: PMC2720697.

5. Prensner JR, Rubin MA, Wei JT, Chinnaiyan AM. Beyond PSA: the next generation of prostate cancer biomarkers. Sci Transl Med. 2012;4(127):127rv3. Epub 2012/03/31. doi: 10.1126/scitranslmed.3003180. PubMed PMID: 22461644; PMCID: PMC3799996.

6. Amling CL, Blute ML, Bergstralh EJ, Seay TM, Slezak J, Zincke H. Long-term hazard of progression after radical prostatectomy for clinically localized prostate cancer: continued risk of biochemical failure after 5 years. J Urol. 2000;164(1):101–5. Epub 2000/06/07. PubMed PMID: 10840432.

7. Han M, Partin AW, Zahurak M, Piantadosi S, Epstein JI, Walsh PC. Biochemical (prostate specific antigen) recurrence probability following radical prostatectomy for clinically localized prostate cancer. J Urol. 2003;169(2):517–23. Epub 2003/01/25. doi: 10.1097/01.ju.0000045749.90353.c7. PubMed PMID: 12544300.

8. Briganti A, Karnes RJ, Gandaglia G, Spahn M, Gontero P, Tosco L, Kneitz B, Chun FK, Zaffuto E, Sun M, Graefen M, Marchioro G, Frohneberg D, Giona S, Karakiewicz PI, Van Poppel H, Montorsi F, Joniau S. Natural history of surgically treated high-risk prostate cancer. Urol Oncol. 2015;33(4):163.e7-13. Epub 2015/02/11. doi: 10.1016/j.urolonc.2014.11.018. PubMed PMID: 25665508.

9. Kim DK, Koo KC, Lee KS, Hah YS, Rha KH, Hong SJ, Chung BH. Time to Disease Recurrence Is a Predictor of Metastasis and Mortality in Patients with High-risk Prostate Cancer Who Achieved Undetectable Prostate-specific Antigen Following Robot-assisted Radical Prostatectomy. J Korean Med Sci. 2018;33(45):e285. Epub 2018/11/08. doi: 10.3346/jkms.2018.33.e285. PubMed PMID: 30402050; PMCID: PMC6209767.

10. Sung H, Ferlay J, Siegel RL, Laversanne M, Soerjomataram I, Jemal A, Bray F. Global Cancer Statistics 2020: GLOBOCAN Estimates of Incidence and Mortality Worldwide for 36 Cancers in 185 Countries. CA Cancer J Clin. 2021;71(3):209–49. Epub 2021/02/05. doi: 10.3322/caac.21660. PubMed PMID: 33538338.

11. Fernandes MC, Yildirim O, Woo S, Vargas HA, Hricak H. The role of MRI in prostate cancer: current and future directions. Magma. 2022;35(4):503–21. Epub 2022/03/17. doi: 10.1007/s10334-022-01006-6. PubMed PMID: 35294642; PMCID: PMC9378354.

12. Turkbey B, Rosenkrantz AB, Haider MA, Padhani AR, Villeirs G, Macura KJ, Tempany CM, Choyke PL, Cornud F, Margolis DJ, Thoeny HC, Verma S, Barentsz J, Weinreb JC. Prostate Imaging Reporting and Data System Version 2.1: 2019 Update of Prostate Imaging Reporting and Data System Version 2. Eur Urol. 2019;76(3):340–51. Epub 2019/03/23. doi: 10.1016/j.eururo.2019.02.033. PubMed PMID: 30898406.

13. Lambin P, Leijenaar RTH, Deist TM, Peerlings J, de Jong EEC, van Timmeren J, Sanduleanu S, Larue R, Even AJG, Jochems A, van Wijk Y, Woodruff H, van Soest J, Lustberg T, Roelofs E, van Elmpt W, Dekker A, Mottaghy FM, Wildberger JE, Walsh S. Radiomics: the bridge between medical imaging and personalized medicine. Nat Rev Clin Oncol. 2017;14(12):749–62. Epub 2017/10/05. doi: 10.1038/nrclinonc.2017.141. PubMed PMID: 28975929.

14. Cutaia G, La Tona G, Comelli A, Vernuccio F, Agnello F, Gagliardo C, Salvaggio L, Quartuccio N, Sturiale L, Stefano A, Calamia M, Arnone G, Midiri M, Salvaggio G. Radiomics and Prostate MRI: Current Role and Future Applications. J Imaging. 2021;7(2). Epub 2021/08/31. doi: 10.3390/jimaging7020034. PubMed PMID: 34460633; PMCID: PMC8321264.

15. Kumar V, Gu Y, Basu S, Berglund A, Eschrich SA, Schabath MB, Forster K, Aerts HJWL, Dekker A, Fenstermacher D, Goldgof DB, Hall LO, Lambin P, Balagurunathan Y, Gatenby RA, Gillies RJ. Radiomics: the process and the challenges. Magnetic Resonance Imaging. 2012;30(9):1234–48. doi: 10.1016/j.mri.2012.06.010.

16. Bartel DP. MicroRNAs: genomics, biogenesis, mechanism, and function. Cell. 2004;116(2):281–97. Epub 2004/01/28. doi: 10.1016/s0092-8674(04)00045-5. PubMed PMID: 14744438.

17. Lenkala D, LaCroix B, Gamazon ER, Geeleher P, Im HK, Huang RS. The impact of microRNA expression on cellular proliferation. Hum Genet. 2014;133(7):931–8. Epub 2014/03/13. doi: 10.1007/s00439-014-1434-4. PubMed PMID: 24609542; PMCID: PMC4677487.

18. Schwarzenbach H. The clinical relevance of circulating, exosomal miRNAs as biomarkers for cancer. Expert Rev Mol Diagn. 2015;15(9):1159–69. Epub 2015/07/24. doi: 10.1586/14737159.2015.1069183. PubMed PMID: 26202667.

19. Barh D, Malhotra R, Ravi B, Sindhurani P. MicroRNA let-7: an emerging next-generation cancer therapeutic. Curr Oncol. 2010;17(1):70–80. Epub 2010/02/25. doi: 10.3747/co.v17i1.356. PubMed PMID: 20179807; PMCID: PMC2826782.

20. Rana S, Valbuena GN, Curry E, Bevan CL, Keun HC. MicroRNAs as biomarkers for prostate cancer prognosis: a systematic review and a systematic reanalysis of public data. Br J Cancer. 2022;126(3):502–13. Epub 2022/01/14. doi: 10.1038/s41416-021-01677-3. PubMed PMID: 35022525; PMCID: PMC8810870.

21. Koppers-Lalic D, Hackenberg M, de Menezes R, Misovic B, Wachalska M, Geldof A, Zini N, de Reijke T, Wurdinger T, Vis A, van Moorselaar J, Pegtel M, Bijnsdorp I. Non-invasive prostate cancer detection by measuring miRNA variants (isomiRs) in urine extracellular vesicles. Oncotarget. 2016;7(16):22566–78. Epub 2016/03/19. doi: 10.18632/oncotarget.8124. PubMed PMID: 26992225; PMCID: PMC5008382.

22. Balzeau J, Menezes MR, Cao S, Hagan JP. The LIN28/let-7 Pathway in Cancer. Front Genet. 2017;8:31. Epub 20170328. doi: 10.3389/fgene.2017.00031. PubMed PMID: 28400788; PMCID: PMC5368188.

23. Gao S, Zhao Z, Wu R, Wu L, Tian X, Zhang Z. MiR-1 inhibits prostate cancer PC3 cells proliferation through the Akt/mTOR signaling pathway by binding to c-Met. Biomed Pharmacother. 2019;109:1406–10. Epub 2018/12/16. doi: 10.1016/j.biopha.2018.10.098. PubMed PMID: 30551391.

24. Gujrati H, Ha S, Mohamed A, Wang BD. MicroRNA-mRNA Regulatory Network Mediates Activation of mTOR and VEGF Signaling in African American Prostate Cancer. Int J Mol Sci. 2022;23(6). Epub 2022/03/26. doi: 10.3390/ijms23062926. PubMed PMID: 35328346; PMCID: PMC8949405.

25. Niture S, Tricoli L, Qi Q, Gadi S, Hayes K, Kumar D. MicroRNA-99b-5p targets mTOR/AR axis, induces autophagy and inhibits prostate cancer cell proliferation. Tumour Biol. 2022;44(1):107–27. Epub 2022/07/12. doi: 10.3233/tub-211568. PubMed PMID: 35811549.

26. Zwanenburg A, Vallières M, Abdalah MA, Aerts H, Andrearczyk V, Apte A, Ashrafinia S, Bakas S, Beukinga RJ, Boellaard R, Bogowicz M, Boldrini L, Buvat I, Cook GJR, Davatzikos C, Depeursinge A, Desseroit MC, Dinapoli N, Dinh CV, Echegaray S, El Naqa I, Fedorov AY, Gatta R, Gillies RJ, Goh V, Götz M, Guckenberger M, Ha SM, Hatt M, Isensee F, Lambin P, Leger S, Leijenaar RTH, Lenkowicz J, Lippert F, Losnegård A, Maier-Hein KH, Morin O, Müller H, Napel S, Nioche C, Orlhac F, Pati S, Pfaehler EAG, Rahmim A, Rao AUK, Scherer J, Siddique MM, Sijtsema NM, Socarras Fernandez J, Spezi E, Steenbakkers R, Tanadini-Lang S, Thorwarth D, Troost EGC, Upadhaya T, Valentini V, van Dijk LV, van Griethuysen J, van Velden FHP, Whybra P, Richter C, Löck S. The Image Biomarker Standardization Initiative: Standardized Quantitative Radiomics for High-Throughput Image-based Phenotyping. Radiology. 2020;295(2):328–38. Epub 2020/03/11. doi: 10.1148/radiol.2020191145. PubMed PMID: 32154773; PMCID: PMC7193906.

27. Lewis BP, Burge CB, Bartel DP. Conserved seed pairing, often flanked by adenosines, indicates that thousands of human genes are microRNA targets. Cell. 2005;120(1):15–20. Epub 2005/01/18. doi: 10.1016/j.cell.2004.12.035. PubMed PMID: 15652477.

28. Chen Y, Wang X. miRDB: an online database for prediction of functional microRNA targets. Nucleic Acids Res. 2020;48(D1):D127–d31. Epub 2019/09/11. doi: 10.1093/nar/gkz757. PubMed PMID: 31504780; PMCID: PMC6943051.

29. Rane JK, Scaravilli M, Ylipää A, Pellacani D, Mann VM, Simms MS, Nykter M, Collins AT, Visakorpi T, Maitland NJ. MicroRNA expression profile of primary prostate cancer stem cells as a source of biomarkers and therapeutic targets. Eur Urol. 2015;67(1):7–10. Epub 2014/09/23. doi: 10.1016/j.eururo.2014.09.005. PubMed PMID: 25234358.

30. Riffo-Campos Á L, Riquelme I, Brebi-Mieville P. Tools for Sequence-Based miRNA Target Prediction: What to Choose? Int J Mol Sci. 2016;17(12). Epub 2016/12/13. doi: 10.3390/ijms17121987. PubMed PMID: 27941681; PMCID: PMC5187787.

31. Balagurunathan Y, Gu Y, Wang H, Kumar V, Grove O, Hawkins S, Kim J, Goldgof DB, Hall LO, Gatenby RA, Gillies RJ. Reproducibility and Prognosis of Quantitative Features Extracted from CT Images. Translational Oncology. 2014;7(1):72–87. doi: 10.1593/tlo.13844.

32. Ruopp MD, Perkins NJ, Whitcomb BW, Schisterman EF. Youden Index and optimal cut-point estimated from observations affected by a lower limit of detection. Biom J. 2008;50(3):419–30. Epub 2008/04/26. doi: 10.1002/bimj.200710415. PubMed PMID: 18435502; PMCID: PMC2515362.

33. Kanehisa M, Furumichi M, Sato Y, Kawashima M, Ishiguro-Watanabe M. KEGG for taxonomy-based analysis of pathways and genomes. Nucleic Acids Res. 2023;51(D1):D587–d92. Epub 2022/10/28. doi: 10.1093/nar/gkac963. PubMed PMID: 36300620; PMCID: PMC9825424.

34. Thomas PD, Ebert D, Muruganujan A, Mushayahama T, Albou LP, Mi H. PANTHER: Making genome-scale phylogenetics accessible to all. Protein Sci. 2022;31(1):8–22. Epub 2021/10/31. doi: 10.1002/pro.4218. PubMed PMID: 34717010; PMCID: PMC8740835.

35. Sherman BT, Hao M, Qiu J, Jiao X, Baseler MW, Lane HC, Imamichi T, Chang W. DAVID: a web server for functional enrichment analysis and functional annotation of gene lists (2021 update). Nucleic Acids Res. 2022;50(W1):W216–w21. Epub 2022/03/25. doi: 10.1093/nar/gkac194. PubMed PMID: 35325185; PMCID: PMC9252805.

36. Litwin MS, Tan HJ. The Diagnosis and Treatment of Prostate Cancer: A Review. Jama. 2017;317(24):2532–42. Epub 2017/06/28. doi: 10.1001/jama.2017.7248. PubMed PMID: 28655021.

37. Šamija I, Fröbe A. GENOMICS OF PROSTATE CANCER: CLINICAL UTILITY AND CHALLENGES. Acta Clin Croat. 2022;61(Suppl 3):86. Epub 2023/03/21. doi: 10.20471/acc.2022.61.s3.13. PubMed PMID: 36938554; PMCID: PMC10022402.

38. Gosselaar C, Roobol MJ, Schröder FH. Prevalence and characteristics of screen-detected prostate carcinomas at low prostate-specific antigen levels: aggressive or insignificant? BJU Int. 2005;95(2):231–7. Epub 2005/01/26. doi: 10.1111/j.1464-410X.2005.05324.x. PubMed PMID: 15667646.

39. Preisser F, Cooperberg MR, Crook J, Feng F, Graefen M, Karakiewicz PI, Klotz L, Montironi R, Nguyen PL, D’Amico AV. Intermediate-risk Prostate Cancer: Stratification and Management. Eur Urol Oncol. 2020;3(3):270–80. Epub 2020/04/19. doi: 10.1016/j.euo.2020.03.002. PubMed PMID: 32303478.

40. Grossman DC, Curry SJ, Owens DK, Bibbins-Domingo K, Caughey AB, Davidson KW, Doubeni CA, Ebell M, Epling JW, Jr., Kemper AR, Krist AH, Kubik M, Landefeld CS, Mangione CM, Silverstein M, Simon MA, Siu AL, Tseng CW. Screening for Prostate Cancer: US Preventive Services Task Force Recommendation Statement. Jama. 2018;319(18):1901–13. Epub 2018/05/26. doi: 10.1001/jama.2018.3710. PubMed PMID: 29801017.

41. Liu W, Xu J. Translation of genomics and epigenomics in prostate cancer: progress and promising directions. Asian J Androl. 2016;18(4):503–4. Epub 2016/06/09. doi: 10.4103/1008-682x.182820. PubMed PMID: 27270344; PMCID: PMC4955169.

42. Jairath NK, Dal Pra A, Vince R, Jr., Dess RT, Jackson WC, Tosoian JJ, McBride SM, Zhao SG, Berlin A, Mahal BA, Kishan AU, Den RB, Freedland SJ, Salami SS, Kaffenberger SD, Pollack A, Tran P, Mehra R, Morgan TM, Weiner AB, Mohamad O, Carroll PR, Cooperberg MR, Karnes RJ, Nguyen PL, Michalski JM, Tward JD, Feng FY, Schaeffer EM, Spratt DE. A Systematic Review of the Evidence for the Decipher Genomic Classifier in Prostate Cancer. Eur Urol. 2021;79(3):374–83. Epub 2020/12/10. doi: 10.1016/j.eururo.2020.11.021. PubMed PMID: 33293078.

43. Zhu C, Hou X, Zhu J, Jiang C, Wei W. Expression of miR-30c and miR-29b in prostate cancer and its diagnostic significance. Oncol Lett. 2018;16(3):3140–4. Epub 2018/08/22. doi: 10.3892/ol.2018.9007. PubMed PMID: 30127906; PMCID: PMC6096223.

44. Duenweg SR, Bobholz SA, Barrett MJ, Lowman AK, Winiarz A, Nath B, Stebbins M, Bukowy J, Iczkowski KA, Jacobsohn KM, Vincent-Sheldon S, LaViolette PS. T2-Weighted MRI Radiomic Features Predict Prostate Cancer Presence and Eventual Biochemical Recurrence. Cancers (Basel). 2023;15(18). Epub 2023/09/28. doi: 10.3390/cancers15184437. PubMed PMID: 37760407; PMCID: PMC10526331.

45. Algohary A, Viswanath S, Shiradkar R, Ghose S, Pahwa S, Moses D, Jambor I, Shnier R, Bohm M, Haynes AM, Brenner P, Delprado W, Thompson J, Pulbrock M, Purysko AS, Verma S, Ponsky L, Stricker P, Madabhushi A. Radiomic features on MRI enable risk categorization of prostate cancer patients on active surveillance: Preliminary findings. J Magn Reson Imaging. 2018. Epub 20180222. doi: 10.1002/jmri.25983. PubMed PMID: 29469937; PMCID: PMC6105554.

46. Fredsøe J, Rasmussen AKI, Mouritzen P, Borre M, Ørntoft T, Sørensen KD. A five-microRNA model (pCaP) for predicting prostate cancer aggressiveness using cell-free urine. Int J Cancer. 2019;145(9):2558–67. Epub 2019/03/25. doi: 10.1002/ijc.32296. PubMed PMID: 30903800.

47. Zhang H, Zhu M, Shan X, Zhou X, Wang T, Zhang J, Tao J, Cheng W, Chen G, Li J, Liu P, Wang Q, Zhu W. A panel of seven-miRNA signature in plasma as potential biomarker for colorectal cancer diagnosis. Gene. 2019;687:246–54. Epub 2018/11/21. doi: 10.1016/j.gene.2018.11.055. PubMed PMID: 30458288.

48. Daugaard I, Sanders KJ, Idica A, Vittayarukskul K, Hamdorf M, Krog JD, Chow R, Jury D, Hansen LL, Hager H, Lamy P, Choi CL, Agalliu D, Zisoulis DG, Pedersen IM. miR-151a induces partial EMT by regulating E-cadherin in NSCLC cells. Oncogenesis. 2017;6(7):e366. Epub 2017/08/02. doi: 10.1038/oncsis.2017.66. PubMed PMID: 28759022; PMCID: PMC5541717.

49. Almeida RS, Costa ESM, Coutinho LL, Garcia Gomes R, Pedrosa F, Massaro JD, Donadi EA, Lucena-Silva N. MicroRNA expression profiles discriminate childhood T-from B-acute lymphoblastic leukemia. Hematol Oncol. 2019;37(1):103–12. Epub 2018/11/06. doi: 10.1002/hon.2567. PubMed PMID: 30393877.

50. Zhang Z, Karthaus WR, Lee YS, Gao VR, Wu C, Russo JW, Liu M, Mota JM, Abida W, Linton E, Lee E, Barnes SD, Chen HA, Mao N, Wongvipat J, Choi D, Chen X, Zhao H, Manova-Todorova K, de Stanchina E, Taplin ME, Balk SP, Rathkopf DE, Gopalan A, Carver BS, Mu P, Jiang X, Watson PA, Sawyers CL. Tumor Microenvironment-Derived NRG1 Promotes Antiandrogen Resistance in Prostate Cancer. Cancer Cell. 2020;38(2):279–96.e9. Epub 2020/07/18. doi: 10.1016/j.ccell.2020.06.005. PubMed PMID: 32679108; PMCID: PMC7472556.

51. Ghosh RD, Pattatheyil A, Roychoudhury S. Functional Landscape of Dysregulated MicroRNAs in Oral Squamous Cell Carcinoma: Clinical Implications. Front Oncol. 2020;10:619. Epub 20200512. doi: 10.3389/fonc.2020.00619. PubMed PMID: 32547936; PMCID: PMC7274490.

52. Zhang S, He Y, Liu C, Li G, Lu S, Jing Q, Chen X, Ma H, Zhang D, Wang Y, Huang D, Tan P, Chen J, Zhang X, Liu Y, Qiu Y. miR-93-5p enhances migration and invasion by targeting RGMB in squamous cell carcinoma of the head and neck. J Cancer. 2020;11(13):3871–81. Epub 20200406. doi: 10.7150/jca.43854. PubMed PMID: 32328191; PMCID: PMC7171485.

53. Greither T, Vorwerk F, Kappler M, Bache M, Taubert H, Kuhnt T, Hey J, Eckert AW. Salivary miR-93 and miR-200a as post-radiotherapy biomarkers in head and neck squamous cell carcinoma. Oncol Rep. 2017;38(2):1268–75. Epub 20170628. doi: 10.3892/or.2017.5764. PubMed PMID: 28677748.

54. Li G, Ren S, Su Z, Liu C, Deng T, Huang D, Tian Y, Qiu Y, Liu Y. Increased expression of miR-93 is associated with poor prognosis in head and neck squamous cell carcinoma. Tumour Biol. 2015;36(5):3949–56. Epub 20150113. doi: 10.1007/s13277-015-3038-6. PubMed PMID: 25578493; PMCID: PMC4445482.

55. Zeljic K, Jovanovic I, Jovanovic J, Magic Z, Stankovic A, Supic G. MicroRNA meta-signature of oral cancer: evidence from a meta-analysis. Ups J Med Sci. 2018;123(1):43–9. Epub 20180226. doi: 10.1080/03009734.2018.1439551. PubMed PMID: 29482431; PMCID: PMC5901467.

56. Yap T, Seers C, Koo K, Cheng L, Vella LJ, Hill AF, Reynolds E, Nastri A, Cirillo N, McCullough M. Non-invasive screening of a microRNA-based dysregulation signature in oral cancer and oral potentially malignant disorders. Oral Oncol. 2019;96:113–20. Epub 20190718. doi: 10.1016/j.oraloncology.2019.07.013. PubMed PMID: 31422202.

57. Meng X, Liang X, Yang S, Wu D, Wang X. A miRNA-7704/IL2RB/AKT feedback loop regulates tumorigenesis and chemoresistance in ovarian cancer. Exp Cell Res. 2024;437(2):114012. Epub 2024/04/03. doi: 10.1016/j.yexcr.2024.114012. PubMed PMID: 38565343.

58. Mahlab-Aviv S, Zohar K, Cohen Y, Peretz AR, Eliyahu T, Linial M, Sperling R. Spliceosome-Associated microRNAs Signify Breast Cancer Cells and Portray Potential Novel Nuclear Targets. Int J Mol Sci. 2020;21(21). Epub 2020/11/05. doi: 10.3390/ijms21218132. PubMed PMID: 33143250; PMCID: PMC7663234.

59. Causin RL, da Silva LS, Evangelista AF, Leal LF, Souza KCB, Pessoa-Pereira D, Matsushita GM, Reis RM, Fregnani J, Marques MMC. MicroRNA Biomarkers of High-Grade Cervical Intraepithelial Neoplasia in Liquid Biopsy. Biomed Res Int. 2021;2021:6650966. Epub 20210413. doi: 10.1155/2021/6650966. PubMed PMID: 33954190; PMCID: PMC8060087.

60. Zhang P, Shao G, Lin X, Liu Y, Yang Z. MiR-338-3p inhibits the growth and invasion of non-small cell lung cancer cells by targeting IRS2. Am J Cancer Res. 2017;7(1):53–63. Epub 2017/01/27. PubMed PMID: 28123847; PMCID: PMC5250680.

61. Liang Y, Xu X, Wang T, Li Y, You W, Fu J, Liu Y, Jin S, Ji Q, Zhao W, Song Q, Li L, Hong T, Huang J, Lyu Z, Ye Q. The EGFR/miR-338-3p/EYA2 axis controls breast tumor growth and lung metastasis. Cell Death Dis. 2017;8(7):e2928. Epub 2017/07/14. doi: 10.1038/cddis.2017.325. PubMed PMID: 28703807; PMCID: PMC5550870.

62. Nie H, Li J, Yang XM, Cao QZ, Feng MX, Xue F, Wei L, Qin W, Gu J, Xia Q, Zhang ZG. Mineralocorticoid receptor suppresses cancer progression and the Warburg effect by modulating the miR-338-3p-PKLR axis in hepatocellular carcinoma. Hepatology. 2015;62(4):1145–59. Epub 2015/06/18. doi: 10.1002/hep.27940. PubMed PMID: 26082033; PMCID: PMC4755033.

63. Sun F, Yu M, Yu J, Liu Z, Zhou X, Liu Y, Ge X, Gao H, Li M, Jiang X, Liu S, Chen X, Guan W. miR-338-3p functions as a tumor suppressor in gastric cancer by targeting PTP1B. Cell Death Dis. 2018;9(5):522. Epub 2018/05/11. doi: 10.1038/s41419-018-0611-0. PubMed PMID: 29743567; PMCID: PMC5943282.

64. Cai C, Zhi Y, Wang K, Zhang P, Ji Z, Xie C, Sun F. CircHIPK3 overexpression accelerates the proliferation and invasion of prostate cancer cells through regulating miRNA-338-3p. Onco Targets Ther. 2019;12:3363–72. Epub 2019/05/24. doi: 10.2147/ott.S196931. PubMed PMID: 31118688; PMCID: PMC6503193.

65. Wang Y, Qin H. miR-338-3p targets RAB23 and suppresses tumorigenicity of prostate cancer cells. Am J Cancer Res. 2018;8(12):2564–74. Epub 2019/01/22. PubMed PMID: 30662812; PMCID: PMC6325485.

66. Huang G, Jiang Z, Zhu W, Wu Z. Exosomal circKDM4A Induces CUL4B to Promote Prostate Cancer Cell Malignancy in a miR-338-3p-Dependent Manner. Biochem Genet. 2023;61(1):390–409. Epub 2022/08/06. doi: 10.1007/s10528-022-10251-2. PubMed PMID: 35930171.

67. Qu F, Zheng J, Gan W, Lian H, He H, Li W, Yuan T, Yang Y, Li X, Ji C, Yan X, Xu L, Guo H. MiR-199a-3p suppresses proliferation and invasion of prostate cancer cells by targeting Smad1. Oncotarget. 2017;8(32):52465–73. Epub 20170418. doi: 10.18632/oncotarget.17191. PubMed PMID: 28881744; PMCID: PMC5581043.

68. Bertacchini J, Mediani L, Beretti F, Guida M, Ghalali A, Brugnoli F, Bertagnolo V, Petricoin E, Poti F, Arioli J, Anselmi L, Bari A, McCubrey J, Martelli AM, Cocco L, Capitani S, Marmiroli S. Clusterin enhances AKT2-mediated motility of normal and cancer prostate cells through a PTEN and PHLPP1 circuit. J Cell Physiol. 2019;234(7):11188–99. Epub 20181122. doi: 10.1002/jcp.27768. PubMed PMID: 30565691.

